# Can accurate demographic information about people who use prescription medications non-medically be derived from Twitter?

**DOI:** 10.1101/2022.04.27.22274390

**Authors:** Yuan-Chi Yang, Mohammed Ali Al-Garadi, Jennifer S. Love, Hannah L. F. Cooper, Jeanmarie Perrone, Abeed Sarker

## Abstract

Traditional surveillance mechanisms for nonmedical prescription medication use (NPMU) involve substantial lags. Social media-based approaches have been proposed for conducting close-to-real-time surveillance, but such methods typically cannot provide fine-grained statistics about subpopulations. We address this gap by developing methods for automatically characterizing a large Twitter NPMU cohort (n=288,562) in terms of age-group, race, and gender. Our methods achieved 0.88 precision (95%-CI: 0.84-0.92) for age-group, 0.90 (95%-CI: 0.85-0.95) for race, and 0.94 accuracy (95%-CI: 0.92-0.97) for gender. We compared the automatically-derived statistics for the NPMU of tranquilizers, stimulants, and opioids from Twitter to statistics reported in traditional sources (*eg*., the National Survey on Drug Use and Health). Our estimates were mostly consistent with the traditional sources, except for age-group-related statistics, likely caused by differences in reporting tendencies and representations in the population. Our study demonstrates that subpopulation-specific estimates about NPMU may be automatically derived from Twitter to obtain early insights.

## INTRODUCTION

Substance use (SU), including non-medical prescription medication use (NPMU), has been a major public health problem in the United States (US) for decades. Overdose deaths due to the SU have steadily increased over the years, regardless of prevention measures.^1^ In 2019, the SU-related overdose death rate increased by 4.3% to 21.6 per 100,000 population,^2^ about 20 times higher than the recorded rate in 1980.^1^ In the 12 months preceding April 2021, over 100,000 SU-related deaths are expected, the highest ever recorded in a 12-month period.^3^ Due to the enormity of the SU epidemic, the US government has announced the deployment of unprecedented resources.^4^

There are also significant disparities related to SU disorder (SUD) and the associated health outcomes. Many recent studies have highlighted the disparities depending on socioeconomic status, race/ethnicity, gender identity/biological sex, community, criminal history, and healthcare coverage.^5-12^ For example, studies have shown that non-Hispanic Blacks and Hispanics are less likely to receive buprenorphine treatment compared to Whites, and women are less likely than men.^11,13-15^ Moreover, non-Hispanic Blacks and American Indians and Alaska Natives (AIAN) experienced the highest increases in the drug overdose mortality rates in 2019 and 2020,^16^ while non-Hispanic Blacks experienced much higher increase of the mortality rates due to stimulants and opioids co-ingestion compared to non-Hispanic Whites.^17^ It has also been reported that people with lower income, living in non-metro urbanized regions, or uninsured are more likely to suffer from SUD.^18^ Multiple disparities may co-exist, and exacerbate the likelihood of SU/SUD. Consequently, non-Hispanic Blacks, Hispanic/Latino persons, AIAN, and Native Hawaiian and Other Pacific Islanders (NHOPI), who also have low insurance coverage rates, face substantial SUD related disparities.^19^ Distinct demographic groups may also have their own unique cultural and historical contexts and norms, consequently increasing the challenges associated with surveillance and response.

A key to effectively tackle the epidemic and alleviate the disparities is to improve surveillance, specifically to accelerate the data curation process to provide timely, actionable insights, and improve decision making.^20,21^ Traditional surveillance approaches and/or sources of data include surveys, such as those conducted by the National Survey on Drug Use and Health (NSDUH),^22^ poison control centers,^23^ hospital data about treatment admissions and discharge,^24^ overdose-related emergency department visits (EDV),^25,26^ and overdose death records.^27^ Such traditional surveillance systems have considerable lags associated with the cycle of data collection, organization and release. For example, the 2020 NSDUH Annual National Report was not available until the end of October 2021. Due to such lags, trends in SU/overdose only detected and understood retrospectively, often after considerable damage has already been done and/or SU patterns have shifted. The lag is particularly problematic since the SU/overdose epidemic has been continuously evolving over the years. For example, the primary contributor of overdose-related deaths in the early 2000s was cocaine, which was later taken over by prescription opioids followed by heroin.^1^ Also, in recent years, there have been notable increases in deaths due to synthetic opioids (*eg*., fentanyl) and psychostimulants (*eg*., methamphetamine),^27-30^ but the current trajectory will only be known after months based on traditional surveillance. Therefore, there is an urgent need for close-to-real-time surveillance system for SU.

To address the shortcomings of traditional approaches, social media have been proposed as potential resources for timely surveillance.^31-33^ Over 220 million Americans (∼70% of the population) use social media, and many discuss health-related topics. There is thus the potential to leverage such patient-generated information for conducting timely surveillance via social media. Social media data, such as that from Twitter, have been shown to contain self-reported information about SU, including NPMU, which can be detected in real-time.^31,34-38^ While social media are promising data sources for obtaining close to real-time insights, data from them are typically massive and noisy, and extracting knowledge from them requires the development of advanced artificial intelligence methods involving natural language processing (NLP) and machine learning. However, research on social media-based SU surveillance is still in its infancy and many developments are required. Motivated lags associated with traditional surveillance systems, we attempted to equip advanced social media-based approaches with effective tools for providing timely understanding of subpopulation-level statistics, which can be used for close-to-real-time surveillance and for studying disparities.

Ideally, SU surveillance data needs to cover the full range of demographics (eg, race, age, gender, geographical area, socioeconomic status), and contain sufficient granularity to observe subtle differences among different demographic groups. Some studies have combined volume of social media information with meta-data such as geolocation and timestamps to derive geolocation-specific SU estimates that correlate with statistics from traditional data sources such as the NSDUH and CDC Wonder database.^18,36-39^ However, to the best of our knowledge, no past study has attempted to estimate demographic information, such as age and gender, from a social media-based cohort. There are multiple reasons why such studies have not been conducted in the past despite their potential utility—the information is not directly available from meta-data, and the proposed estimation methods in the literature do not have adequate accuracy or granularity.^40-50^ This poses a barrier to conducting fine-grained, subpopulation-specific research using such data—a clear disadvantage compared to the NSDUH and other traditional sources. Methods for accurately and automatically estimating the distributions of key demographic features in social media subscriber cohorts can enable such fine-grained subpopulation-level analyses and comparisons.

In this paper, we describe the development and validation of methods for automatically estimating demographic distributions (age-group, gender, and race) in a Twitter cohort consisting of subscribers who self-reported NPMU. We integrated our developed novel methods to establish a data-centric cohort characterization pipeline, and applied the pipeline on the Twitter NPMU cohort. To validate our pipeline, we compared the distributions estimated from Twitter to those reported in traditional sources [NSDUH 2019^18^ and Nationwide Emergency Department Sample (NEDS)^51^] for prescription stimulants, tranquilizers, and opioid pain relievers.

## RESULTS

### Twitter nonmedical use cohort

We collected tweets mentioning prescription medications and detected self-reported NPMU using a supervised classification system.^35^ Posts were collected from March 6, 2018 to April 30, 2021. Our system detected 482,902 NPMU-indicating tweets and extracted their authors’ meta-data, including post history, if available. In this manner, we collected 288,562 Twitter subscribers’ posts (NPMU cohort).

### Gender, age, and race distribution

The gender, age, and race proportions for Twitter subscribers, estimated from the 2018 Twitter Survey conducted by the Pew Research Center,^52^ and those reported in the US Census^18^ are shown in Figure 1. The estimated gender and race proportions from the two sources are comparable, while the age proportions are substantially different. Compared to the US Census data, Twitter has marginally lower proportions of females (4% less) and Whites (1.5% less), and more Hispanics (1.5% more), which may be explained by the overcount of Whites and undercount of Hispanics in Census.^53^ The closeness of the proportions from the two sources suggest that Twitter-based estimates specific to gender and race may be representative of the country’s population. In contrast, in terms of age, Twitter has an overrepresentation of younger people compared to the census estimates. Specifically, the proportion of people in the 18-25 group is approximately 10% higher, and the proportion for the 55+ years group is 20% lower on Twitter compared to the census estimates. The overrepresentation of younger people on Twitter, and social media in general, is a well-known phenomenon.

**Figure 1.**
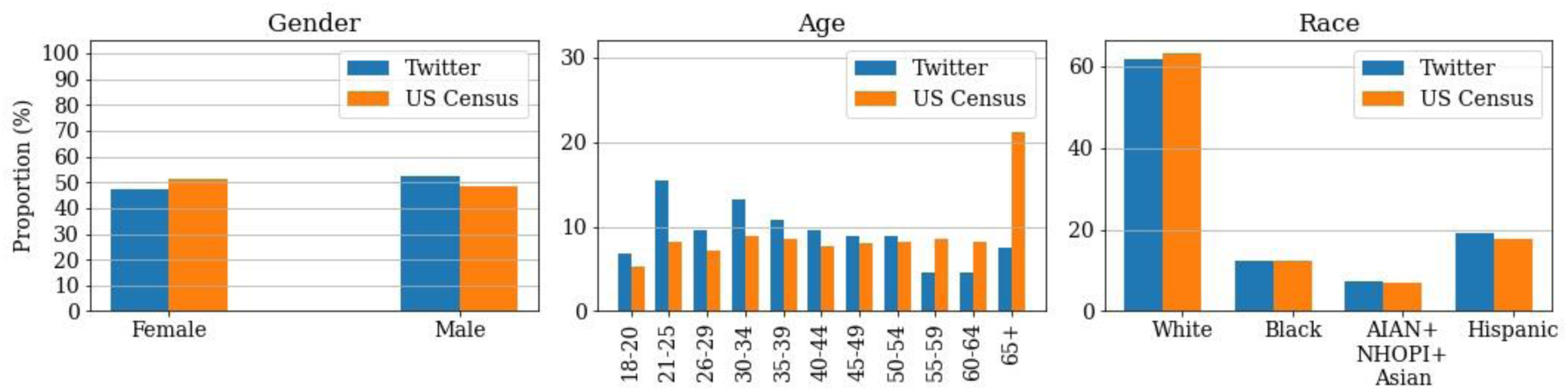
Gender, age, and race proportions estimated from Twitter and those reported in US census.

### Gender distribution estimates

The estimated gender proportions from Twitter data and the gender proportions from traditional sources, including NSDUH and NEDS, are given in Figure 2 (details in Supplement Table S1). The three categories of medications included were opioid pain relievers, tranquilizers, and stimulants. For NPMU of tranquilizers, the estimated Twitter proportions are within the 95% confidence intervals of the proportions reported in the NSDUH. For stimulants, the Twitter proportion estimate for females is slightly higher than the NSDUH reported number (∼5%). For opioids, the estimated proportions are substantially different between Twitter and the NSDUH (∼10%). Specifically, the proportion for females from Twitter is lower than the NSDUH estimates. Interestingly, however, we found that the numbers reported by the NSDUH also differ in terms of proportions from the opioid-related EDVs reported in NEDS, but the estimates from the latter are very close to the Twitter proportions (no significant difference). This suggests that estimates derived from Twitter may be more predictive of overdose-related events rather than nonmedical use for this category.

**Figure 2.**
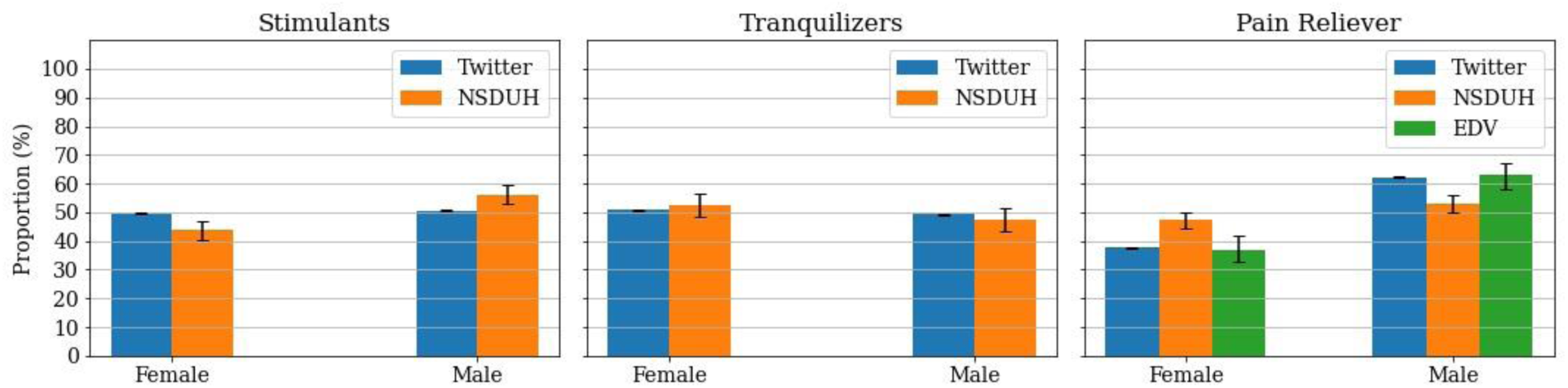
Gender distributions for nonmedical prescription medication use estimated from Twitter and those reported in the NSDUH. For opioid pain relievers, the gender distribution of overdose-related emergency medicine visits is also provided.

**Figure 3.**
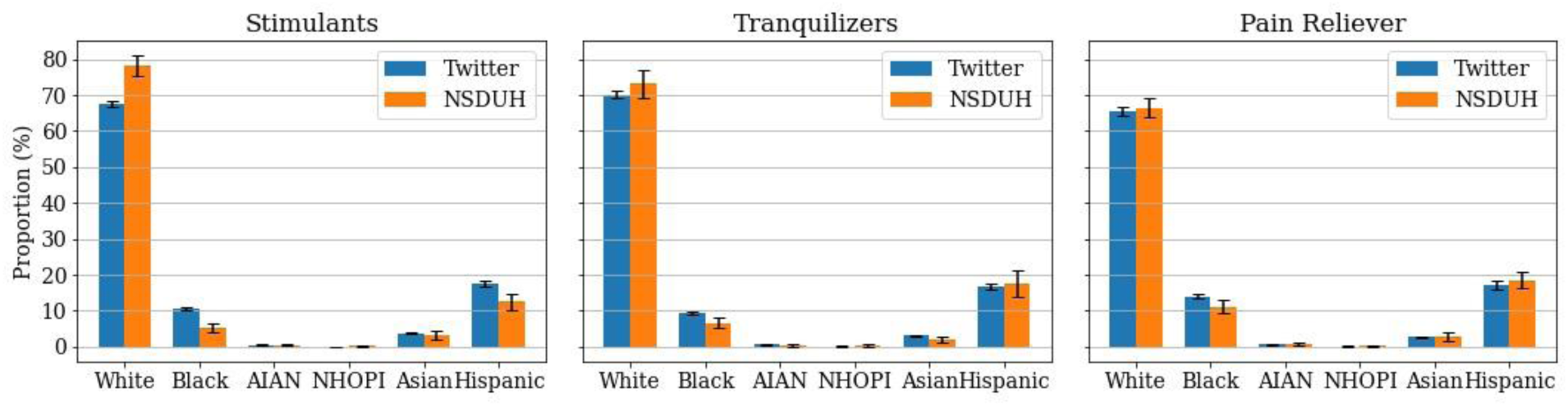
Race distributions for nonmedical prescription medication use estimated from Twitter and those reported in the NSDUH.

**Figure 4.**
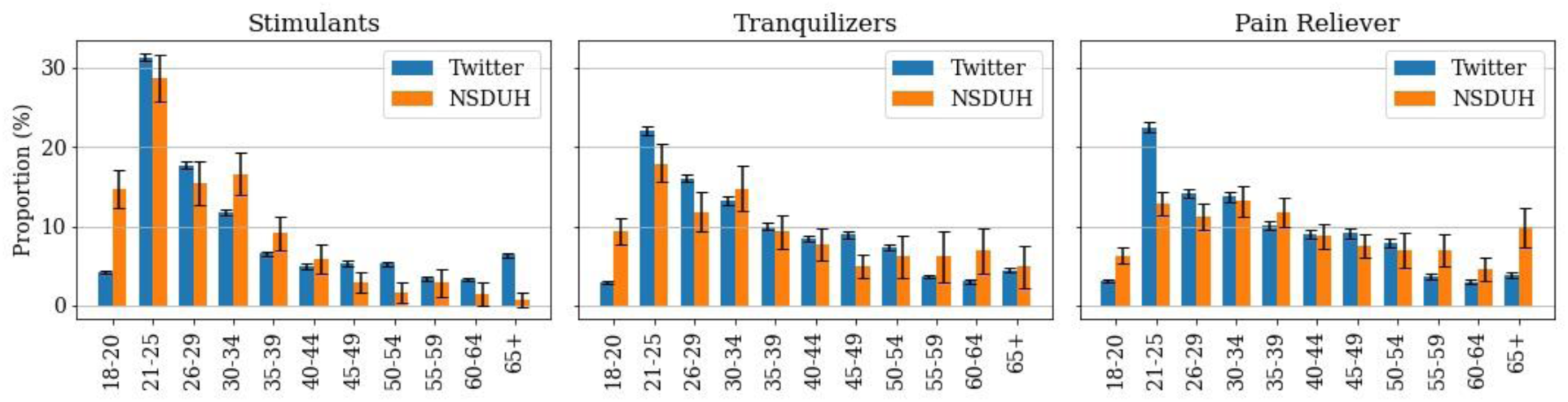
Age-group distributions for nonmedical prescription medication use estimated from Twitter and those reported in the NSDUH.

### Race distribution estimates

The estimated race proportions from Twitter and the proportions from the NSDUH are shown in Figure (details in Supplement Table S2). The estimated distributions are similar between the two data sources, and each proportion from Twitter is either within or close to the 95% confidence interval of the corresponding proportion reported in the NSDUH. For all medication categories, the majority of people who reported nonmedical use are White, followed by Hispanic and Black. Asians who reported nonmedical use only represent about 4% or less of the cohort; American Indian and Alaska Native (AIAN) and the Native Hawaiian or Other Pacific Islander (NHOPI) groups each represent less than 1% of the cohort. Importantly, the Twitter data had representation from all of the minority races. The most prominent differences between Twitter and the NSDUH are for the White and Hispanic stimulant groups, and for Blacks across all medications (Twitter estimates are higher than NSDUH).

### Age-group distribution estimates

The estimated age-group proportions from Twitter data and the proportions from the NSDUH are shown in Figure (details in Supplement Table S3). For most age-groups, the Twitter and the NSDUH estimates are similar. The most prominent differences are for young adults (18-20 and 21-25) and the elderly (65+). The estimated proportions from Twitter are consistently lower for the 18-20 group and higher for the 21-25 group. For the 65+ group, the estimated Twitter proportion is higher for stimulants, similar for tranquilizers, and lower for pain relievers compared to the NSDUH numbers. For opioid pain relievers, the estimated Twitter proportion for the 21-25 group is approximately 10% higher and for the 65+ group 6% lower compared to the NSDUH. For tranquilizers, the estimated Twitter proportion for the 18-20 group is approximately 6% lower compared to the NSDUH, but the proportions for the 65+ group are not significantly different. For simulants, the estimated Twitter proportion for the 18-20 group is approximately 10% lower and for the 65+ group 6% higher compared to the NSDUH.

## DISCUSSION

To the best of our knowledge, our current work is the first to automatically estimate the distribution of demographic characteristics in a large Twitter cohort—in this case a cohort of subscribers who self-reported NPMU—and compare the automatically-obtained distributions with those reported in traditional sources. Our experiments validate that most of the estimates derived from Twitter are consistent with those reported in traditional sources, such as the NSDUH and NEDS. The major differences were in the age-group-based estimates, specifically for young adults (18-20 and 21-25) and the elderly (65+).

The NSDUH is conducted as a survey among noninstitutionalized population in the US and thus is limited by the respondents’ truthfulness and exclusion of individuals in hospitals, prisons, or even treatment centers.^54^ It is reported that the respondents tend to under-report or over-report on surveys, and this tendency is influenced by their demographics including gender, race, or age.^55-59^ For example, among surveyed Cocaine users, African American, young adults (18-30), and female are more inclined to under-report.^55^ Though Twitter data also relies on individual users’ truthfulness and willingness to share, we suspect that the default anonymity of Twitter accounts renders the Twitter data suffer less demographic-wise under-reporting than the NSDUH and, thus, might be better suited for analyzing subpopulation differences than the NSDUH. We speculate that the closeness of the gender and race distributions for Twitter subscribers and the US population, as depicted in Figure 1, is a key reason for the Twitter-based estimates to be similar to the NSDUH, while the differences might be explained by the under/over-reporting tendency. For example, the under-reporting tendency of females on Cocaine might help explain the apparent overestimation of female stimulant users on Twitter, and the under-reporting tendency of African American might be crucial to understand the overestimation of African American users for all three medication categories on Twitter.^55^ Similarly, the different tendencies of under/over-reporting and Twitter usage among age groups might contribute to the differences in the age-wise estimates. Also, though the Twitter data is limited to those who have internet access, it may capture a certain portion of the institutionalized or before/after their institutionalized periods. It is even possible that Twitter is less biased against the incarcerated Black population than the NSDUH.

### Evaluation of performance as a surveillance system

Our pipeline is advantageous in its timeliness, flexibility, simplicity, and stability. The data collection can be done near-real time and continuously on a personal desktop with internet connection while requiring minimal human supervision. Instead of structured answers to questionnaires, our collected data contain salient unstructured text information, allowing data mining with research questions evolving over time. For example, the collected tweets contain information regarding how and why the authors are using the medications. To collect similar information from surveys usually requires incorporating prior knowledge into question design, but that is not necessary for our pipeline. However, since the data is usually massive, the data analysis is typically done using NLP and machine-learning methods. The advantage is that, once the tailored scripts are developed, it can often be run automatically and on the fly. To expand the pipeline (eg, to collect users of illicit drugs), it would only require a university informatics team to dedicate a few months from initial exploration to operational prototypes. These advantages of our system fit nicely into the CDC’s Data Modernization Initiative, Priority 2 - Accelerate Data into Action to Improve Decision-Making and Protect Health.

However, the data quality, acceptability, sensitivity, the predicted value positive, and representativeness for our pipeline are inevitably limited by Twitter’s user base and Twitter users’ willingness to share their information publicly and their truthfulness. Demographic information is often not disclosed and we have no method to validate the users’ claims. Notably, our data does not rely on memory as much as surveys, as the time leap is limited to the time of non-medical use to the time of posting. Also, we speculated that the users might be more truthful in their self-disclosure because they have more power to choose what they are willing to share publicly. Therefore, though our method does have disadvantages, we believe it is an important complimentary surveillance system with high potential.

### Related work

Our work is not the first that aims to detect demographic information from social media data, although it is the first to develop and apply such methods for characterizing a specific cohort. Past studies have proposed cohort characterization methods, including gender,^40-43^ age,^42,44-46^ and race.^47-50^ Typically, these pipelines comprised supervised classification methods, and used subscribers’ meta-data including names, usernames, bio, past tweets, or even images as features. Among these, the gender detection methods were reported to be the most accurate, with classifiers achieving accuracies above 94%.^60^ In contrast, race and age estimation pipelines proposed in past research had not obtained high accuracies. They also often do not provide enough granularity. The race estimation pipelines reported in the literature usually focus only on four categories (White, Black, Asian, and Hispanic/Latino) or less, leaving out AIAN and NHOPI,^47-50^ although AIAN has the highest overdose mortality rate among all race group.^16,61^ For age detection, the groupings often do not match those defined in the NSDUH, making comparisons impossible.^44-46^ Though re-grouping is possible for a few methods, they were not developed based on Twitter and thus may have limited applicability.^42^ In our work, we developed age and race estimation pipelines with high precision and fine granularity (11 for age and six for race) based on our Twitter data. Due to the paucity of annotated datasets, we applied a search-based approach that employs text pattern matching for detecting self-disclosed age and race. Because there is no gold-standard for the negative case (*ie*., subscribers who have not self-disclosed their age or race using the specified pattern), we focused on improving precision while maintaining acceptable retrieval rate. Precision is preferred over recall (*ie*., some cohort members will be missed) since the number of Twitter cohort members is large and growing over time, so obtaining sufficient numbers of people from each category is not a bottleneck.

### Potential applications and future work

There are several aspects of our automatic pipeline that can be improved in the future. First, we can improve the performance of the classifier that detects the self-disclosures of NPMU. One potential route is to examine if the classifier underperforms for any demographic group due to different self-disclosure behaviors, and then fine-tune the classifier accordingly. Second, we may make our findings more reliable by further improving the age group and race characterization methods. Three potential directions include (i) annotating more tweets matched by the text patterns, (ii) enriching the set of the text patterns, and/or (iii) replacing the rule-based method with a machine-learning based classifier. Third, the pipeline can be extended to illicit substances, including opioids such as heroin, and stimulants such as methamphetamine. Collecting a cohort of people who self-report the use of illicit substances is an important and natural extension to our current work, as Literature suggests that people who report NPMU may also be exposed to illicit substances.^62^ Fourth, our pipeline provides access to targeted minority groups, which is of great interest to the communities and several government agencies.^63-65^ Exacerbated by the war on drugs and inequalities in enforcement and incarceration, some of these groups are extremely hard to reach by traditional means. As Twitter features the default anonymity, our pipeline can reach, for example, Black males, to create an unprecedentedly large cohort and to understand their unique challenges and potential effective interventions. Furthermore, our pipeline allows longitudinal study, in comparison to the cross-sectional nature of NSDUH.

### Limitations

Our work is largely limited by the data source. The demographic distribution of Twitter subscribers is different than the US population, especially for age groups. Though we adjust the estimates according to the 2018 Twitter survey, it only partially solves the issue. For people who do not use Twitter or do not discuss their non-medical use, we have no other means to collect their data and, thus, there may be groups of people that are not represented in our analyses. Also, social media data is noisy and may contain false information (*eg*., fake races or genders), which we have no alternative approach to verify. Additionally, tweets are short (limited by 280 characters) and consist of colloquial words, posing significant limitations on the classifier development, and thus the performance of the overall pipeline.

Our proposed methods *cannot* and *should not* be used for identifying the age group, race or gender identity of individual Twitter subscribers. Their performances are not perfect so there is no guarantee that they will not incorrectly characterize a single subscriber. Our methods essentially *estimate* the distributions of demographic information within a given *cohort*. Due to the large size of the cohort, we anticipate that the small numbers of incorrect characterizations are eclipsed.

Finally, an important limitation of our current work is that it excludes certain segments of the population who have been underrepresented in past research—such as those with non-binary gender identities. Our current work could not achieve such level of granularity due to lack of available data. The promising findings from our current work is the first step and our planned future work includes improving the inclusive of our analysis (*eg*., by inclusion of non-binary population, the uninsured, and those unreachable via traditional means).

## METHODS

### Twitter nonmedical use cohort

Twitter data was collected through a data processing pipeline that we have described in past research.^34,35^ The components of the pipeline include collecting publicly available streaming data about prescription medications, classifying the data, and then retrospectively collecting the metadata of subscribers who are detected to self-report NPMU. We collected English tweets mentioning at least one of over 20 PMs (including generic and trade names, street names, and common misspellings)^66^ that have the potential for nonmedical use (see Supplement Table S4).^34^ We developed annotation guidelines with our domain expert (JP), and annotated a subset consisting of 16,443 tweets into four categories: *non-medical use, consumption, information*, or *non-relevant* (see Supplement Table S5 and S6 for annotation details).^34^ We used the annotated data to train machine-learning classifiers, and the best performing classifier (based-on RoBERTa-Large, with an accuracy of 82.3%) was deployed in our data collection pipeline. Our cohort consists of tweets that were signaled as *non-medical use* by the machine-learning classifier.

### Gender distribution estimates

The genders of the Twitter subscribers were estimated based on the meta-1 classifier in our prior work.^60^ The genders are treated in a binary framework (excluding those with non-binary gender identities due to lack of available data) and should be interpreted as how users represent themselves online and thus are closer to the user’s gender identities than biological sexes. We developed the classifier based on gender-labeled datasets made available by Liu & Ruths^41^ and Volkova et al.^67^ In total, we were able to retrieve the meta-data of 67,181 subscribers, consisting of 35,812 (53.3%) females and 31,369 (46.7%) males, which we used to develop the pipeline. We validated the performance on people who use prescription medications nonmedically from Twitter on a set of 412 subscribers whose genders were identified using their linked, public Facebook profiles. The classifier achieved accuracy of 94.4% (95%-Confidence Intervals: 92.0%-96.6%).

### Age distribution estimates

A rule-based approach, which searches for text patterns that are self-disclosures about the subscribers’ ages, was employed to detect age-groups. Further rules are applied to resolve inconsistencies among detected information. The pattern matching is done using regular expressions. Sample text patterns include “*(\d\d) birthday to me*” or “*i’m (\d\d)*” where “*\d*” denotes digits (0-9). We also constructed a filter to remove irrelevant statements that are not associated with ages, such as “*I’m 20 weeks pregnant*.” The pipeline was developed based on a set of 2,000 subscribers, among which 1,540 tweets from 609 subscribers matched with the text patterns and were annotated. The annotation agreement based on an overlapping set of 346 subscribers (952 tweets) is 89.3% with Cohen’s *K*=0.89 (95.8% with Cohen’s *K*=0.96 on tweets). The test accuracy is 0.88 (95% CI: 0.84-0.92) [0.90 (95% CI: 0.86-0.94) if allowed 1 year age discrepancy] on the subscribers who have matched tweets (referred as precision in the text) and 0.93 (95% CI: 0.90-0.95) on the matched tweets.

### Race distribution estimates

The race estimation module is similar to the age estimation one and applies rules and patterns. Relevant expressions indicating race are searched using regular expressions. Example text patterns include “*i’m (black)*” or “*i’m (white)*”. We also constructed a filter to remove irrelevant statements such as “*I’m black salmon*.” The pipeline was developed based on a set of 4,000 tweets, among which 1,124 tweets from 578 subscribers matched with the text patterns and were annotated. The annotation agreement based on an overlapping set of 293 subscribers (533 tweets) is 87.7% with Cohen’s *K*=0.78 (94.0% with Cohen’s *K*=0.88 on tweets). The test accuracy of the pipeline is 0.90 (95% CI: 0.85-0.95) on the subscribers who have matched tweets (referred as precision in the text) [0.94 (95% CI: 0.91-0.97) on the matched tweets]. Since the pipeline is not designed for subscribers who have more than 1 race (“more” in the 2018 Twitter Survey^68^ and the NSDUH^18^), we did not include those subscribers when reporting the results and comparing with the references.

### Baseline for Twitter subscribers’ demographics

We established the baseline for the US Twitter subscribers based on the 2018 Twitter Survey conducted by Pew Research Center.^68^ We focused on Twitter subscribers who have at least used Twitter once a week and calculated the proportions of the targeted demographics (eg., age, gender, race) among these subscribers. We noted that the survey was conducted only among the adults (aged 18+) and the Asian, AIAN, and NHOPI races were grouped into “others.”

### Baseline for Twitter age and race characterization

We established the baseline for Twitter Age and Race characterization based on a data set of 156,368 general Twitter subscribers. We first collected streaming tweets with stopwords in nltk package as keywords on Aug 27, 2021. Data collection using stopwords is an attempt to collect a random set of Twitter subscribers. We then collected the subscribers’ metadata and applied the age and race pipelines. Our objective was to calibrate our pipeline by estimating how many of the subscribers within certain age or race groups actually self-disclosed their age and race on Twitter and were captured by our pipeline. The estimated rates (of detection/self-disclosure) were then used as weights to normalize the age/race proportions obtained from the NPMU dataset.

### Calculation of the proportions for PWUS based on Twitter data

For each subscriber’s characteristics (gender, age, and race), we used the number of Twitter subscribers in each category, inferred by the corresponding pipelines, to estimate the proportion. For age and race, this calculation was limited on the Twitter subscribers whose age or race can be inferred. The proportions were further normalized via the rates of detection as:

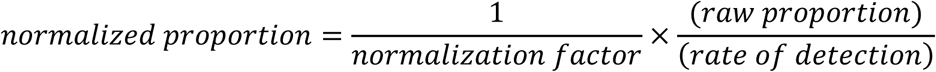

For example, if 10% of the Twitter subscribers are black and only 1% of the random Twitter subscribers disclose that they are black, then we can estimate that roughly 1 in 10 black Twitter subscribers self-disclose their race (rate of detection). Then if we captured 100 people who reported the nonmedical use of stimulants and disclosed that they are black, we infer that roughly 1000 people who use stimulants captured in our pipeline are black and use this number to calculate the normalized proportion. For the race proportions, since AIAN, NHOPI, and Asian were combined into “others” category in the 2018 Twitter Survey, we only calibrated for the “others” category as a whole and assumed that their relative proportions are the same as obtained using the race characterization pipeline.

### Calculation of proportions for the NSDUH data

We established the baseline for the NSDUH/US Census based on Table 12.1A (age) and Table 12.2A (gender and race) in the 2019 NSDUH. We calculated the gender, age, and race proportion through the estimated “Numbers in Thousands.” For age, we used the “Total (2019)” column. For the age and gender, we used the “Age 12+ (2019)” column. For the gender and race for people who report NPMU, we again used the “Age 12+ (2019)” column on Table 1.47A (stimulants), Table 1.53A (tranquilizers), and table 1.44A (pain relievers). For age, we used the “Misuse in the past Year (2019)” column on Table 1.14A (stimulants), Table 1.16A (tranquilizers), and table 1.13A (pain relievers).

### Calculation of proportions for the NEDS data

We calculated the gender proportion of the EDV by using the “No.” column for all opioid poisoning on the Supplemental Table 2C in the Annual Surveillance Report of Drug-related Risks and Outcomes.^51^ The weighted estimates provided in the table are from the NEDS 2016.

### Estimation of 95% confidence intervals

For the Twitter data and the test performance of the pipelines, the confidence intervals are estimated via bootstrapping. For the NSDUH and NEDS data, the 95% confidence intervals are estimated using simulation. For each category, we approximate the distribution as a normal distribution with the reported number as mean and the standard error as standard deviation. We then repeatedly sampled the joint distribution for all the categories in the targeted demographics and calculated the proportions, assuming each category is independent of each other. The 95% confidence intervals were then constructed using the 0.025 quantile and the 0.975 quantile within the list of proportions of the given category. For the NEDS data, the standard errors for estimated numbers (“No.” column) were estimated through the standard errors (“SE” column) for rates (“Rate” column) as 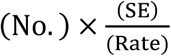.

## Data Availability

The datasets generated during and/or analysed during the current study are not publicly available due to Twitter API's use terms and privacy concern but are available from the corresponding author on reasonable request.

## AUTHOR CONTRIBUTIONS

YY conducted and directed the machine learning experiments, evaluations and data analyses, with assistance from MAA and AS. AS provided supervision for full study. JSL, HLFC, and JP provided toxicology and public health domain expertise for interpreting the results. YY drafted the manuscript and all authors contributed to the final manuscript.

## ACKNOWLEDGEMENTS

The authors thank the support from the National Institute of Health and National Institute of Drug Abuse.

