## Supplementary Materials for "Can accurate demographic information about people who use prescription medications non-medically be derived from Twitter?"

Jennifer S. Love, MD^2^

Hannah L. F. Cooper, ScD^3^

Jeanmarie Perrone, MD^4^

Abeed Sarker, PhD^1,5^

^1^Department of Biomedical Informatics, School of Medicine, Emory University, Atlanta, GA, United States;

^2^Department of Emergency Medicine, Icahn School of Medicine at Mount Sinai, New York, NY, United States;

^3^Department of Behavioral Sciences & Health Education, Rollins School of Public Health, Emory University, Atlanta, GA, United States;

^4^Department of Emergency Medicine, Perelman School of Medicine, University of Pennsylvania, Philadelphia, PA, United States;

^5^Department of Biomedical Engineering, Georgia Institute of Technology and Emory University, Atlanta, GA, United States;

*Corresponding author

Postal address: 101 Woodruff Circle, 4th Floor East, Atlanta, GA 30322

Table S1. The gender proportion of baseline (Twitter), and the NMUPM and NMU for stimulants, tranquilizers, and opioids, along with 95% confidence intervals

| Dataset | Twitter (95%-CI) (%) | | NSDUH (95%-CI) (%) | | EDV (95%-CI) (%) | |
| --- | --- | --- | --- | --- | --- | --- |
|  | F | M | F | M | F | M |
| baseline | 47.5 | 52.5 | 51.5 | 48.5 | - | - |
| NMU stimulant | 49.5^†^  (49.8-49.2) | 50.5^†^  (50.8-50.2) | 43.8  (47.1-40.4) | 56.2  (59.6-52.9) | - | - |
| NMU tranquilizer | 50.8  (51.1-50.5) | 49.2  (49.5-48.9) | 52.5  (56.6-48.5) | 47.5  (51.5-43.4) | - | - |
| NMU opioids | 37.7^†^  (38.1-37.3) | 62.3^†^  (62.7-61.9) | 47.2  (50.1-44.2) | 52.8  (55.8-49.9) | 37.0  (41.8-32.7) | 63.0  (67.3-58.2) |

^†^ The Twitter estimates lie outside the 95 confidence intervals from NSDUH 2019

Table S2. The race proportion baseline (Twitter), and the NMUPM and NMU for stimulants, tranquilizers, and Pain Relievers, along with 95% confidence intervals

| Dataset | Twitter (95%-CI) (%) | | | | | | NSDUH (95%-CI) (%) | | | | | |
| --- | --- | --- | --- | --- | --- | --- | --- | --- | --- | --- | --- | --- |
|  | W* | B | AI | N | A | H | W | B | AI | N | A | H |
| baseline^‡^ | 61.6 | 12.2 | 7.2 | | | 19.0 | 63.2 | 12.3 | 0.6 | 0.4 | 5.9 | 17.6 |
| NMU stimulant | 67.5^†^  (68.3-66.7) | 10.7^†^  (11.1-10.3) | 0.5 (0.6 - 0.5) | 0.1 (0.1 - 0.1) | 3.8 (4.1 - 3.6) | 17.4^†^ (18.2-16.6) | 78.3  (81.1-75.5) | 5.3  (6.6-4.0) | 0.5  (0.8-0.2) | 0.2  (0.4-0.0) | 3.3  (4.6-2.1) | 12.4  (14.8-10.1) |
| NMU tranquilizer | 70.2  (71.1-69.2) | 9.2^†^ (9.6-8.8) | 0.6  (0.7-0.5) | 0.1  (0.1-0.1) | 3.2^†^ (3.4-2.9) | 16.8  (17.7-15.9) | 73.1  (76.9-69.3) | 6.7  (8.3-5.1) | 0.3  (0.6-0.1) | 0.4  (0.8-0.0*) | 1.9  (2.8-1.0) | 17.7  (21.3-13.9) |
| NMU opioids | 65.5  (66.7-64.3) | 14.0^†^  (14.6-13.4 | 0.7  (0.8-0.5) | 0.1  (0.2-0.0) | 2.7  (3.0-2.4) | 17.1  (18.3-15.9) | 66.5  (69.3-63.7) | 11.2  (13.0-9.4) | 0.8  (1.2-0.5) | 0.2  (0.5-0.0) | 2.8  (3.9-1.6) | 18.4  (20.7-16.2) |

^†^ The Twitter estimates lie outside the 95 confidence intervals from NSDUH 2019

^‡^The 2018 Twitter Survey combine Asian, AINA, and NHOPI as others.

* W: white non-Hispanic; B: black non-Hispanic; AI: American Indian and Alaska Native, N: Native Hawaiian or Other Pacific Islander; A: Asian; M: Two or More Races; H Hispanic or Latino

^§^ The simulation gives a negative lower limit on 95% confidence interval calculation

Table S3. The age proportion of baseline (Twitter), and the NMUPM and NMU for stimulants, tranquilizers, and opioids, along with 95% confidence intervals

| Dataset | Twitter (95%-CI) (%) | | | | | | | | | | | NSDUH (95%-CI) (%) | | | | | | | | | | |
| --- | --- | --- | --- | --- | --- | --- | --- | --- | --- | --- | --- | --- | --- | --- | --- | --- | --- | --- | --- | --- | --- | --- |
|  | 18-20 | 21-25 | 26-29 | 30-34 | 35-39 | 40-44 | 45-49 | 50-54 | 55-59 | 60-64 | 65+ | 18-20 | 21-25 | 26-29 | 30-34 | 35-39 | 40-44 | 45-49 | 50-54 | 55-59 | 60-64 | 65+ |
| baseline | 6.9 | 15.5 | 9.6 | 13.2 | 10.7 | 9.6 | 8.9 | 9.0 | 4.5 | 4.6 | 7.4 | 5.3 | 8.2 | 7.2 | 8.9 | 8.6 | 7.7 | 8.1 | 8.2 | 8.5 | 8.2 | 21.1 |
| NMU stimulant | 4.2^†^ (4.4-4.0) | 31.3 (31.8-30.8) | 17.7  (18.1-17.3) | 11.7  (12.1-11.3) | 6.6^†^  (6.9-6.2) | 4.9  (5.2-4.7) | 5.3^†^  (5.6-4.9) | 5.3^†^  (5.6-4.9) | 3.4  (3.6-3.2) | 3.3^†^  (3.5-3.1) | 6.4^†^  (6.7-6.1) | 14.8  (17.2-12.4) | 28.6  (31.6-25.7) | 15.5  (18.3-12.6) | 16.6  (19.3-13.9) | 9.1  (11.2-7.0) | 5.8  (7.7-4.0) | 2.9  (4.1-1.6) | 1.6  (2.9-0.3) | 2.8  (4.6-1.1) | 1.5  (2.9-0.0^§^) | 0.8  (1.6-0.0^§^) |
| NMU tranquilizer | 2.9^†^  (3.0-2.8) | 22.1^†^  (22.5-21.6) | 16.0^†^  (16.5-15.6) | 13.2  (13.7-12.8) | 10.0  (10.4-9.6) | 8.4  (8.8-8.1) | 9.0^†^  (9.4-8.5) | 7.3  (7.7-6.9) | 3.6  (3.9-3.4) | 3.0^†^  (3.2-2.8) | 4.4  (4.7-4.1) | 9.3  (11.0-7.6) | 17.9  (20.4-15.6) | 11.8  (14.3-9.3) | 14.7  (17.6-11.9) | 9.3  (11.4-7.2) | 7.7  (9.8-5.7) | 5.0  (6.5-3.5) | 6.2  (8.9-3.5) | 6.2  (9.4-2.9) | 7.0  (9.8-4.0) | 4.9  (7.5-2.3) |
| NMU opioids | 3.1^†^  (3.3-2.9) | 22.5^†^  (23.1-21.8) | 14.1  (14.7-13.5) | 13.7^†^  (14.3-13.1) | 10.1  (10.7-9.6) | 9.0  (9.5-8.4) | 9.1  (9.8-8.5) | 7.9 (8.4-7.4) | 3.6^†^  (4.0-3.3) | 3.0^†^  (3.2-2.7) | 3.9^†^  (4.2-3.5) | 6.3  (7.3-5.3) | 12.9  (14.4-11.4) | 11.2  (12.9-9.5) | 13.2  (15.0-11.3) | 11.7  (13.5-9.9) | 8.8  (10.3-7.2) | 7.6  (9.1-6.1) | 7.1  (9.2-4.8) | 7.0  (9.0-4.9) | 4.6  (6.1-3.1) | 9.8  (12.2-7.4) |

^†^ The Twitter estimates lie outside the 95 confidence intervals from NSDUH 2019

^§^ The simulation gives a negative lower limit on 95% confidence interval calculation

Table S4: Prescription medications used in data collection for the Toxicovigilance dataset (Dataset-2)

| Medication Category | Medications |
| --- | --- |
| Tranquilizers | Ativan, Valium, Clonazepam, Alprazolam, Xanax, Lorazepam, Diazepam, Klonopin |
| Stimulants | Methylphenidate, Ritalin, Vyvanse, Lisdexamphetamine, Adderall |
| Pain relievers (that contains opioids) | Oxycodone, Vicodin, Morphine, Oxycontin, Percocet, Zohydro, Tramadol, Buprenorphine, Methadone, Suboxone, Hydrocodone, Avinza, Conzip, Dolophine |
| Others | Abilify, Zyprexa, Quetiapine, Seroquel, Risperidone, Aripiprazole, Risperdal, Saphris, Olanzapine, Asenapine, Gabapentin, Pregabalin, Lyrica, Gralise, Neurontin |

Table S5: A brief description of annotation guidelines for tweets that indicate non-medical use of Prescription Medication. Extracted from O’Connor et al.^1^

i. The tweet explicitly states that the user has taken or is going to take the medication to *experience certain feelings* (ie, to get high) or that the user *experienced certain feelings in the past*.
ii. The tweet expresses that the user *has or is going to coingest a medication with other prescription medications or illicit drugs or alcohol or coffee* (or other substances).
iii. The tweet expresses a *mechanism of intake* that is typically associated with abuse or misuse.

Table S6: Examples of tweets that indicate non-medical user and tweets that don't indicate non-medical user. Extracted from Table 2 in Sarker et al.^2^

| **Tweets that indicate non-medical use** | **Tweets that don’t indicate non-medical use** |
| --- | --- |
| popped Adderall tonight hahahah let’s finish this 100 page paper | Seroquel is prescribed. i use valerian root sometimes too. mostly i don’t sleep |
| an oxycodone high from snorting lasts for one hour, if it is swallowed, your looking at three hour high | a prescription for adderall should come with my college acceptance paper speaking of oxycodone .. i need to take mine. This pain is ridiculous |
